# COVID-19 DEATH TOLL: THE ROLE OF THE NATION’S ECONOMIC DEVELOPMENT

**DOI:** 10.1101/2020.07.18.20156778

**Authors:** Gabriel Chodick, Clara Weil

## Abstract

**Background:** Several months into the novel coronavirus disease (COVID-19) pandemic, there is a limited understanding of the underlying country-specific factors associated with COVID-19 spread and mortality. This study aims to investigate the role of nations’ economic development in the death toll associated with COVID-19 in Europe and Israel.

**Methods:** Number of COVID-19 cases, deaths per million, and case fatality rate (CFR) in Israel and 39 countries in Europe were described across quintiles of gross domestic product (GDP) per capita. The association between GDP per capita and COVID-19 incidence, mortality, and CFR was investigated using generalized linear modeling adjusting for the proportion of elderly and density of the population.

**Results:** In countries belonging to the three lower GDP quintiles, COVID-19 incidence rates per million (range 708-1134) were substantially lower compared to countries in the fourth (3939) and fifth (3476) quintiles. Major differences were also calculated in COVID-19 mortality rates per million (25-31 vs. 222-268). There was no significant (p=0.19) differences in CFR between GDP quintiles (range: 2.79-7.62%).

**Conclusions:** COVID-19 had a greater toll in more developed nations. Though comparisons are limited by differences in testing, reporting and lockdown policies, this association likely reflects increased spread from trade and tourism in wealthier countries, whereas limited health system capacity and lack of treatment and vaccination options contributed to higher than expected CFR in wealthier countries. This unique situation will probably encourage the stronger economies to invest the required financial capacity to respond to and recover from the current crisis.

## INTRODUCTION

The novel coronavirus SARS-CoV-2 (severe acute respiratory syndrome coronavirus 2) causing coronavirus disease (COVID-19) was first identified in Wuhan, China, in December 2019. It is transmitted quite efficiently since substantial transmission is possible by patients with mild symptoms or no symptoms. The illness is characterized by a relatively long, clinically mild phase that can last 5-9 days before symptoms are severe enough to seek medical attention^1^. Since viral shedding peaks at the start of the illness when symptoms are mild, the early phase of the disease is the most critical risk period for community transmission^2^. It has been recently estimated that 44% of secondary cases are infected before the onset of symptoms in the primary case^3^.

The first months of 2020 saw rapid spread of COVID-19 outside of China, leading the World Health Organization (WHO) to declare a pandemic by 11 March 2020. To date, there is limited understanding of the underlying factors associated with COVID-19 spread and mortality, and research is ongoing around the world. Italy, for example, may have the oldest population in Europe, but this explains only in part its high COVID-19 death toll. Early Chinese and Italian experiences underscored the challenges of cross-country comparisons of COVID-19 case and mortality rates, as countries differ in their proportion of elderly citizens, testing policies, and reporting of cases and deaths ^4^. Cross-country comparisons have also explored potential correlations with Bacille Calmette-Guérin vaccination policies, with conflicting reports ^5^.

Israel (which belongs to the WHO European region) reported its first COVID-19 cases on 21 February 2020 and established early and strict lockdown measures to contain the spread. Early trajectories of incidence in Israel were similar to European countries such as Austria, while COVID-19 mortality remained relatively low compared.^6^ In this study, we aim to investigate the role of nations’ economic development in explaining differences in the death toll associated with COVID-19 in Europe and Israel.

## METHODS

The number of reported COVID-19 cases and deaths in Israel and European countries was obtained from Worldmeter databases^7^ on May 28, 2020. To increase correlation stability we have excluded countries of small populations limiting the standard error of mortality rate to less than 10 per million. A total of 40 countries were included in the analysis. Cumulative COVID-19 cases (incidence) and deaths were reported per million population and case fatality rate (CFR) was calculated as the number of reported COVID-19 deaths per 1000 cases.

GDP per capita (in purchasing power parity USD) ^8^ was used as a proxy for the countries’ economic development.COVID-19 incidence, mortality, and CFR were described across quintiles of gross domestic product (GDP) per capita; Wald Chi-Square tests were used for pairwise comparisons between the lowest quintile and higher quintiles. Generalized linear modeling (assuming a normal distribution with an identity link function) was also used to investigate the association of GDP per capita with COVID-19 incidence, mortality and CFR after adjusting for the proportion of elderly (≥65y) ^9^ and population density (people per sq. km of land area)^9^. All p-values were two-sided, and p-values < 0.05 were considered statistically significant. Analyses were performed using R 3.5.1 and IBM-SPSS version 25

## RESULTS

The analysis included 39 countries, with GDP quintiles classified into: lowest (<$23154), second ($23155-31964, e.g. Portugal), third ($31965-$39499, e.g. Italy and Israel), forth ($39500-$51936 e.g. Austria, Belgium, UK, Spain), and highest ($51937 or above $, e.g. Netherlands) (Appendix table 1).

### COVID19 incidence

The spread of the virus in this region was highly associated with economic wealth (**Figure 1**). Significant difference (Wald Chi-Square p=0.001) in COVID-19 incidence rate per million were calculated between national GDP levels. Pairwise comparisons in the GLM model taking into account national elderly rate and crowding indicate that incidence in the lowest GDP per capita quintile (median= 979 per million) was comparable (p>0.60) to the second (780) or third (708) quintiles, it was significantly (p=0.001) lower as compared to the fourth (3939, p=0.002) and the fifth (3476, p=0.003) quintiles.

**Figure 1.**
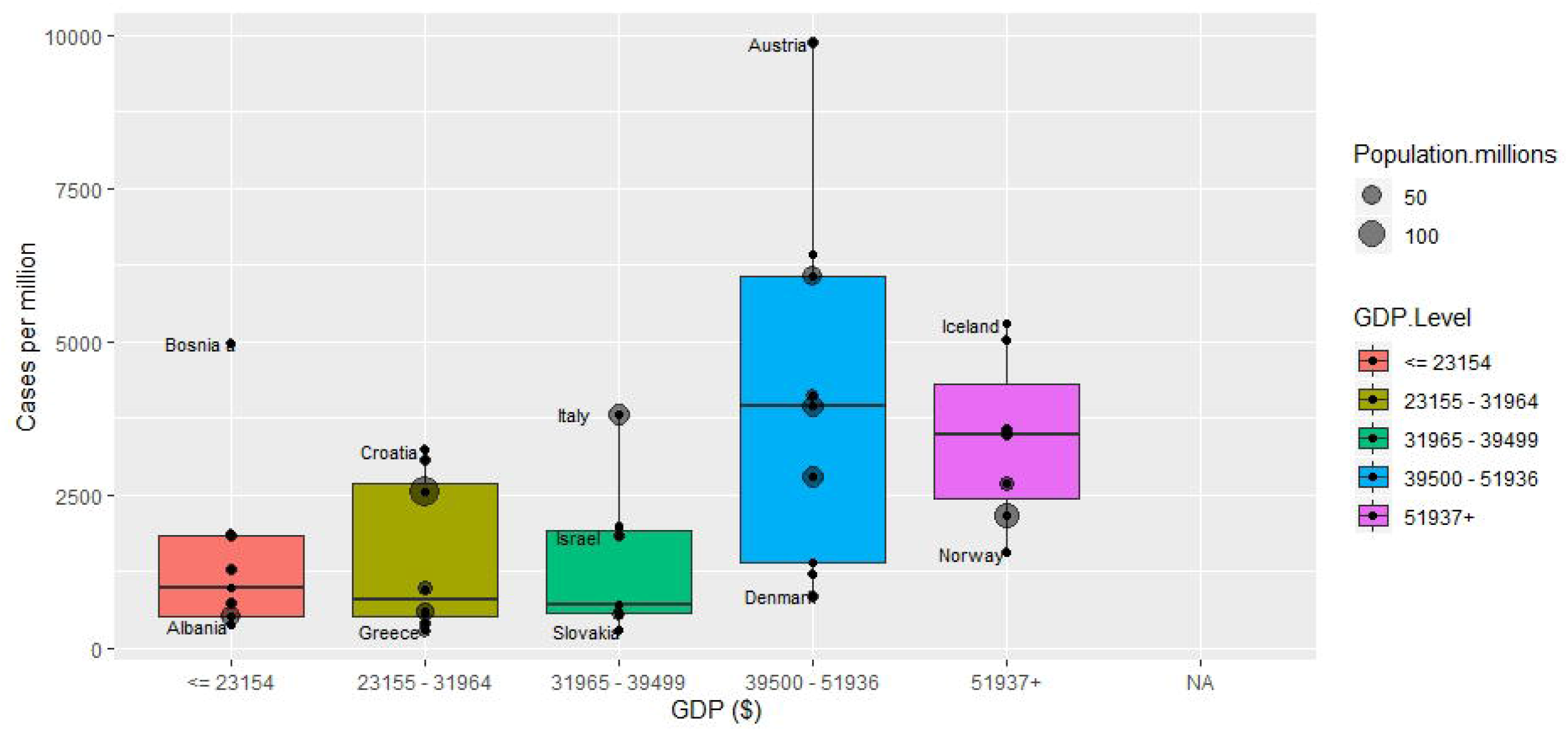

### COVID19 case-fatality ratio

COVID-19 case fatality ratio per national GDP quintile is given in Figure 2. In the GLM model there was no significant (p=0.304) differences between GDP quintiles, with median CFR ranging between 2.79% to 7.62%

**Figure 2.**
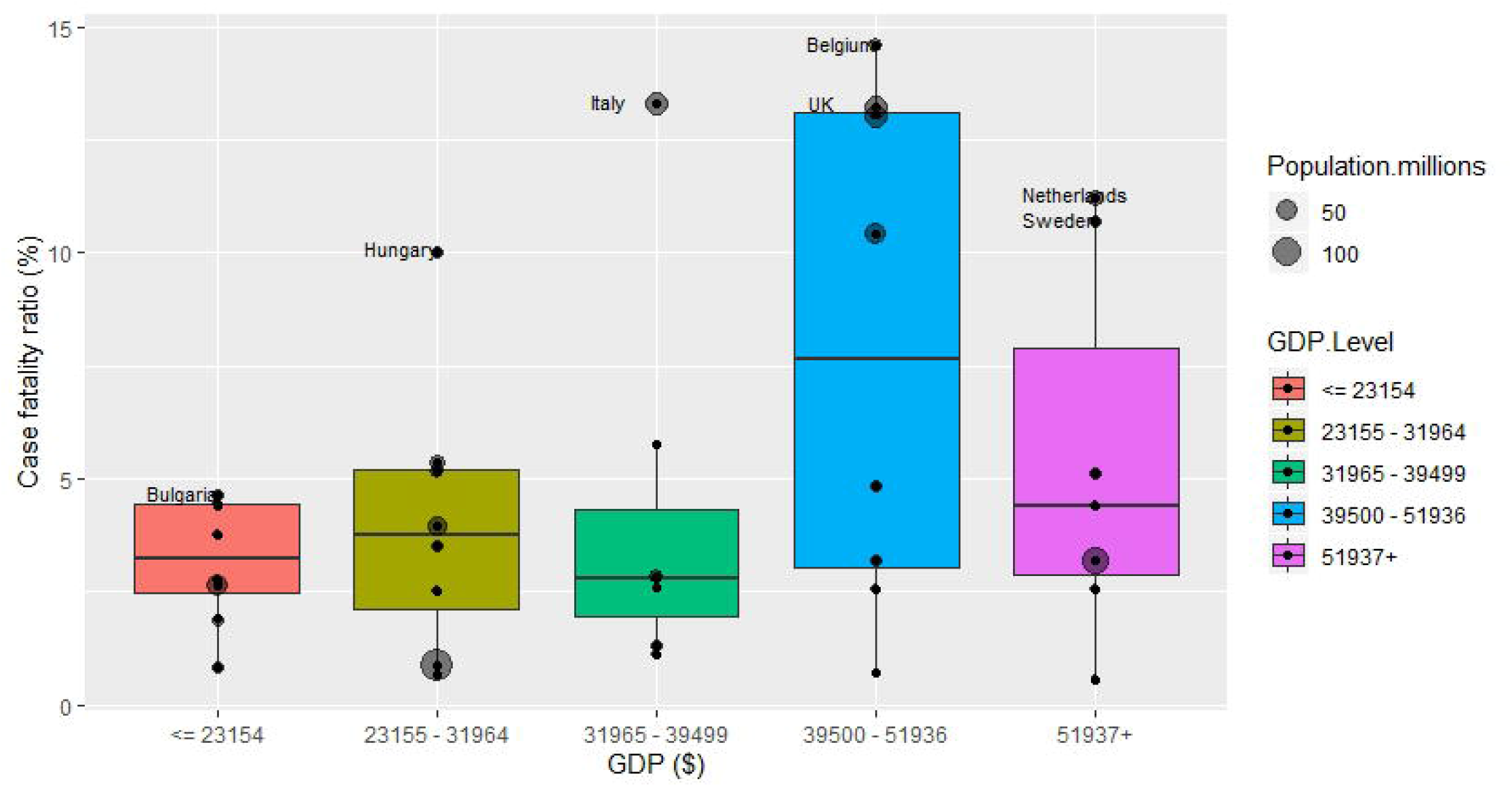

### COVID19 mortality rate

Countries with higher GDP per capita had significantly higher COVID-19 mortality rates (**Figure 3**). Pairwise comparisons adjusted for proportion of elderly and density indicate that while no significant (p>0.4) difference was observed in mortality rates per million between the lowest quintiles (median, 23) as compared to the second (27) or third quintiles (31), but substantially lower compared to countries of the fourth (268, p=0.002) and the fifth (222, p=0.04) quintiles.

**Figure 3.**
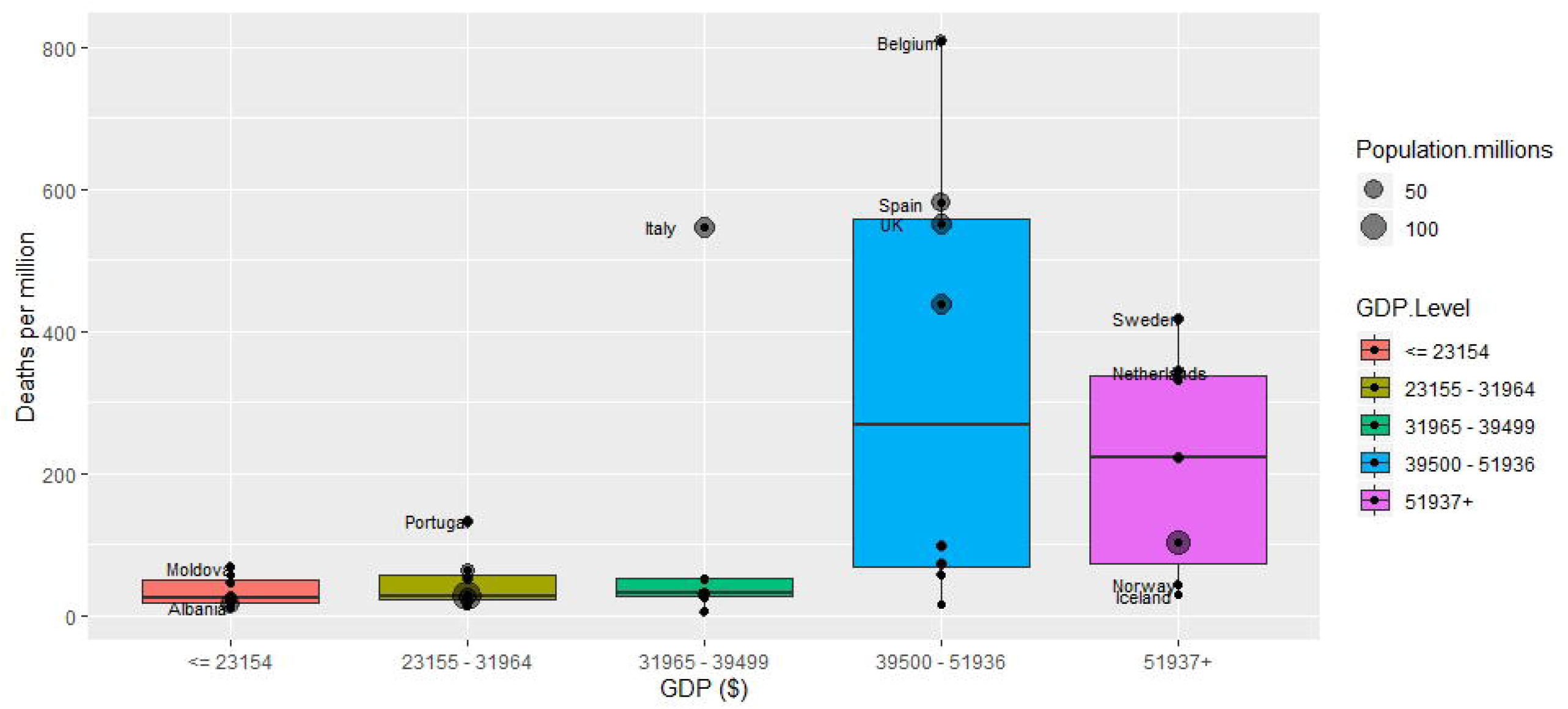

## DISCUSSION

In the European region, wealthier countries with higher GDP per capita reported higher incidence rates of COVID-19, but had no advantage in treating diagnosed cases as indicated by relatedly high CFR Consequently, COVID-19 had a significantly higher death toll in high GDP nations. The higher incidence in the more developed countries of the continent can be explained by the greater flow of cases from China for business and tourism at the early stage of the outbreak and the open borders policy. As a result, the local population in these countries was probably more likely to be exposed to a larger number of cases important from China. European countries with highest COVID-19 mortality rates also include those countries with the top busiest airport (exc. Germany) and passenger traffic : the UK, France, the Netherlands, and Spain^9^.

In contrast to influenza, COVID-19 has currently no effective treatment or vaccination. Thus, wealthier countries showed no advantage in patient survival as indicated by case fatality ratio (CFR). The higher than expected CFR calculated in countries with high incidence, such as Italy and Belgium can be explained by the crisis of the intensive care systems. This has led to a strong association between COVID-19 mortality and GDP per capita.

Clearly, there are many differences between countries in the reporting quality and methods, availability and access to COVID-19 tests. Underreporting of cases in nursing homes, in particularly, may contribute to variation across the region. Estimates of excess deaths suggest that countries such as Belgium – whose high death toll includes victims who had coronavirus-like symptoms but did not test positive – appears to capture almost all the estimated excess. In contrast, Austria’s reported deaths captured only people who had tested positive, accounting for an estimated 57% of excess deaths.^10,11^

Differences in household structures, intergenerational ties, and care of the elderly have also been correlated with the COVID-19 CFR and may contribute to differences at the global level. In addition, generalizability of these findings outside the European region may be limited, as variability in factors such as income inequality, health system capacity^13^ and policies may play a more important role. Further research is needed to investigate the relationship between economic development indices with COVID-19 mortality in other regions.

However, the conclusion from this analysis is that, quite rarely, COVID-19 had a greater toll in more developed nations. This unique situation will probably encourage the stronger economies to invest the required financial capacity to develop effective vaccines and therapies for COVID-19 to quickly re-emerge from the ashes of the current crisis.

## Data Availability

All data were collected from publically avaliable resources

---

**Table 1:** Countries indcluded in the analysis, by GDP quintile (2019)^8^

| **GDP per capita level** |  |  |  |  |
| --- | --- | --- | --- | --- |
| **Lowest** | **2<sup>nd</sup>** | **3<sup>rd</sup>** | **4<sup>th</sup>** | **Highest** |
| Ukraine | Russia | Slovenia | UK | Germany |
| Serbia | Romania | Slovakia | Spain | Iceland |
| North Macedonia | Portugal | Lithuania | Malta | Ireland |
| Montenegro | Poland | Italy | Luxemburg | Netherlands |
| Moldova | Latvia | Israel | France | Norway |
| Bulgaria | Hungary | Estonia | Finland | Sweden |
| Bosnia and Herzegovina | Greece | Czech Rep. | Denmark | Switzerland |
| Belarus | Croatia |  | Belgium |  |
| Albania |  |  | Austria |  |

